# Dynamic Estimation of Epidemiological Parameters of COVID-19 Outbreak and Effects of Interventions on Its Spread

**DOI:** 10.1101/2020.04.01.20050310

**Authors:** Hongzhe Zhang, Xiaohang Zhao, Kexin Yin, Yiren Yan, Wei Qian, Bintong Chen, Xiao Fang

**Author notes:** These first authors contributed equally to the work (Hongzhe Zhang, Xiaohang Zhao, Kexin Yin). Correspondence to (Xiao Fang), (Bintong Chen), (Wei Qian).

## Abstract

A key challenge for estimating the epidemiological parameters of the COVID-19 outbreak in Wuhan is the discrepancy between the officially reported number of infections and the true number of infections. A common approach to tackling the challenge is to use the number of infections exported from Wuhan to infer the true number in the city. This approach can only provide a static estimate of the epidemiological parameters before Wuhan lockdown on January 23, 2020, because there are almost no exported cases thereafter. Here, we propose a method to dynamically estimate the epidemiological parameters of the COVID-19 outbreak in Wuhan by recovering true numbers of infections from day-to-day official numbers. Using the method, we provide a comprehensive retrospection on how the disease had progressed in Wuhan from January 19 to March 5, 2020. Particularly, we estimate that the outbreak sizes by January 23 and March 5 were 11,239 [95% CI 4,794–22,372] and 124,506 [95% CI 69,526–265,113], respectively. The effective reproduction number attained its maximum on January 24 (3.42 [95% CI 3.34–3.50]) and became less than 1 from February 7 (0.76 [95% CI 0.65–0.92]). We also estimate the effects of two major government interventions on the spread of COVID-19 in Wuhan. In particular, transportation suspension and large scale hospitalization respectively prevented 33,719 and 90,072 people from getting infected in the nine-day time period right after its implementation.

## Introduction

A novel coronavirus has quickly spread across China and penetrated into many other countries since December 2019^1^. As of April 6, 2020, the virus has infected 83,005 individuals in China and 1,210,956 individuals globally according to WHO reports^2^. An essential step to contain or slow the outbreak of COVID-19 (i.e., the disease caused by the novel coronavirus) is to uncover its epidemiological parameters over time so that we can analyze the effect of different interventions on its spread^3^. Toward that end, a number of studies have attempted to estimate its epidemiological parameters such as the number of infected cases and the reproduction number^1,4,5,6,7,8,9^. A key challenge for these studies is that the officially reported number of infections (hereafter referred to as the official number) could be much lower than the true number of infections, especially in the early stage of the pandemic and at the center of the pandemic in China, the city of Wuhan^10^. This under-reporting problem could be attributed to many factors, such as insufficient amount of virus test kits and the shortage of hospital beds.

A common approach to tackling the under-reporting problem is to use the official number of infected cases exported from Wuhan to infer the true number of infections within Wuhan, assuming that, outside Wuhan, the official number is close to the true number^4,5,7^. For example, Wu et al. ^5^ use the number of cases exported from Wuhan internationally to infer the true number of infections in Wuhan whereas Cao et al. ^4^ employ the official number of cases exported from Wuhan domestically. This approach can only provide a static estimate of the epidemiological parameters before January 23, 2020, because there are almost no exported cases from Wuhan after the Wuhan lockdown effective January 23, 2020^11^. However, the epidemiological parameters of the COVID-19 are dynamic, partly because of various interventions over time. It is therefore imperative to estimate the epidemiological parameters of the COVID-19 outbreak dynamically and beyond January 23, 2020.

Here, we solve the under-reporting problem from a distinctive perspective. Rather than relying on cases exported from Wuhan, we propose a method to dynamically estimate the epidemiological parameters of the COVID-19 outbreak in Wuhan over time by transforming day-do-day official numbers of infections. Specifically, we propose a Bayesian estimation method that seamlessly integrates a epidemic model characterizing the spread mechanism of the disease and a salient transformation approach, coupled with prior knowledge on key parameters of the epidemic model. Our proposed method has the following distinguishing features compared to existing methods. First, we tackle the under-reporting problem by proposing a straightforward yet effective transformation approach to adjust for potential discrepancies between official and true numbers to give better overall picture for the scope of the COVID-19 outbreak, thereby more reliably quantifying its key epidemiological parameters. Second, our approach conveniently incorporates the fast evolving knowledge from new COVID-19 literature to generate well-justified and more refined parameter estimation results with uncertainty quantification. Furthermore, the temporal dynamic estimation over time keeps track of the evolving disease spread in response to interventions and holds the promise of objectively monitoring and evaluating effectiveness of various containment measures. Our analysis uncovers and demonstrates the evolution of the COVID-19 outbreak in Wuhan from January 19, 2020 to March 5, 2020. In particular, for every day in this period, we apply the proposed method to estimate the effective reproduction number as well as true numbers of infections, such as the cumulative number of infected cases and the number of actively infected but not quarantined cases. Our proposed method also produces daily under-reporting factors, which indicate the degree of discrepancies between official and true numbers. Finally, using the dynamic epidemiological parameters estimated by our analysis, we evaluate the effects of two major interventions on the spread of COVID-19 in Wuhan.

## Results

### Outbreak Size in Wuhan

Using our approach detailed in the Method section, we estimated the true cumulative number of infections in Wuhan by each day for the period between January 19, 2020 and March 5, 2020. The input to our method is the cumulative number of infections in Wuhan by January 18, 2020 estimated in Imai et al. ^12^, whose baseline estimate is 4,000 with a 95% confidence interval [1,700–7,800]. Fig. 1 plots the true cumulative number of infections estimated by our method in a dotted blue line, in comparison to its respective official number reported by the government (solid blue line). As shown, the gap between these two curves is significant, especially at the beginning of the observation period measured by percentage. Such marked difference is partly attributable to the lack of testing and treatment capacities, especially at the beginning of the outbreak. In particular, we estimated that the true cumulative numbers of infections in Wuhan by January 23, 2020 (date of Wuhan lockdown) and March 5, 2020 were 11,239 [95% CI 4,794–22,372] and 124,506 [95% CI 69,526–265,113], respectively. In comparison, their respective official numbers were 495 and 49,797. We also provide our estimated true cumulative number of infections in Wuhan by each day in the observation period (Supplementary Table 1).

**Fig. 1.**
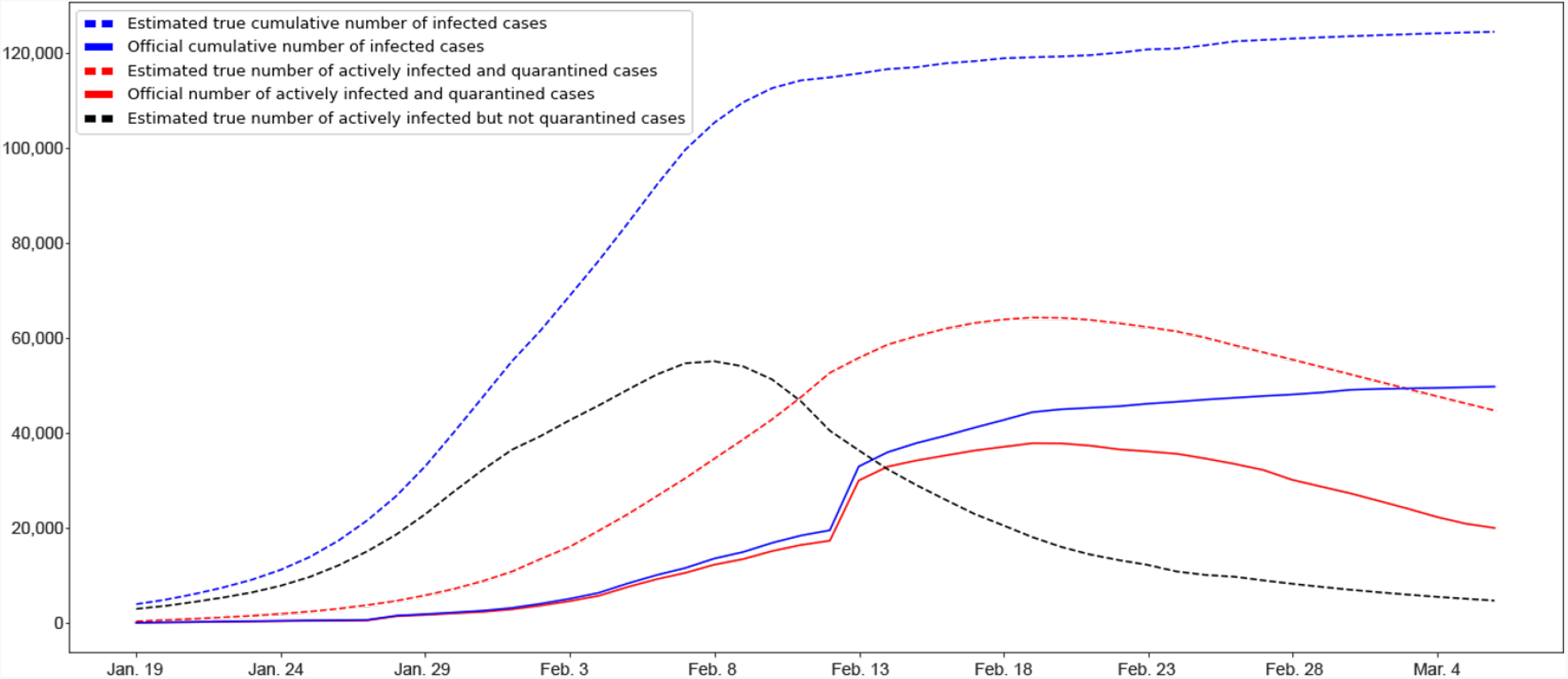
Estimated and Official Numbers of Infections in Wuhan. We plot true numbers of infections estimated by our method in dotted lines and official numbers of infections in solid lines. The dotted blue line presents the estimated true cumulative number of infected cases or the outbreak size, whereas its respective official number is given in the solid blue line **(Blue)**. The estimated outbreak size in Wuhan by March 5, 2020 was 124,506 [95% Cl 69,526-265,113], The dotted red line gives the estimated true number of actively infected and quarantined cases, whereas its official counterpart is given in the solid red line **(Red)**. The estimated true number of actively infected but not quarantined cases is given in the dotted black line **(Black)**.

Fig. 1 also presents the estimated true number of actively infected and quarantined cases by each day in the observation period (dotted red line) and its respective official number (solid red line). The former is computed by our method, which estimates the actual number of actively infected cases who are quarantined effectively, whereas the latter typically counts those actively infected and currently quarantined at hospitals. By March 5, 2020, our estimated true number of actively infected and quarantined cases was 44,778 [95% CI 24,049–112,697] whereas its official counterpart was 20,049. The gap between these two curves represents the number of actively infected people who are effectively quarantined but fail to be included in the government statistics. Many of these infected people could not be tested or officially admitted to hospital, but nevertheless conducted effective self-quarantine at home or other isolated places.

The last curve in the figure shows the estimated true number of actively infected but not quarantined cases by each day in the observation period (dotted black line). It refers to the number of actively infected people who are not quarantined at all (e.g., non-symptomatic infected cases^13^) or not quarantined effectively (i.e., still being able to infect others). These infected people were not recorded by government reports either. Hence, we do not have the official number of actively infected but not quarantined cases. As shown, the estimated true number of actively infected but not quarantined cases peaked on February 7, 2020 (55,139 [95% CI 24,204–118,273]) and then started to decline. This decline was due to the operation of a number of new hospitals and a major COVID-19 testing facility^14^. As a result, many of those actively infected but not quarantined got tested and hospitalized.

### Evolution of the Effective Reproduction Number

Fig. 2 plots the evolution of the effective reproduction number *R* in Wuhan from January 19, 2020 to February 24, 2020, with the shaded area representing the 95% credible interval. As discussed in the Method section, *R* is estimated using a rolling-window approach with 10-day window size. Therefore, *R* of day *t* indicates the transmissibility of COVID-19 in Wuhan over the time window of [*t, t* + 10]. Three major government measures illustrated in the figure include Wuhan lockdown effective January 23, 2020, which stopped all inner-city and inter-city public transportations, vehicle ban effective January 26, 2020, which suspended all non-essential taxi, ride-hailing operation and private vehicle services, and large scale hospitalization beginning on February 5, 2020, which tested and hospitalized a large number of infected people due to added testing and treatment capacities. As shown in the figure, *R* of January 19, 2020 was 3.11 [95% CI 2.93–3.40]. It then climbed up and attained its maximum on January 24, 2020, which was 3.42 [95% CI 3.34–3.50]. This initial surge could be partly attributed to increased gathering and friend visiting during the period of the Chinese Spring Festival. The effective reproduction number *R* declined from January 24, 2020. This could be due to the two government measures that suspended transportation in Wuhan and subsequently reduced the average contact rate among Wuhan residents. The large scale hospitalization started on February 5 further reduced *R* and it became less than 1 from February 7, 2020 (0.76 [95% CI 0.65–0.92]).

**Fig. 2.**
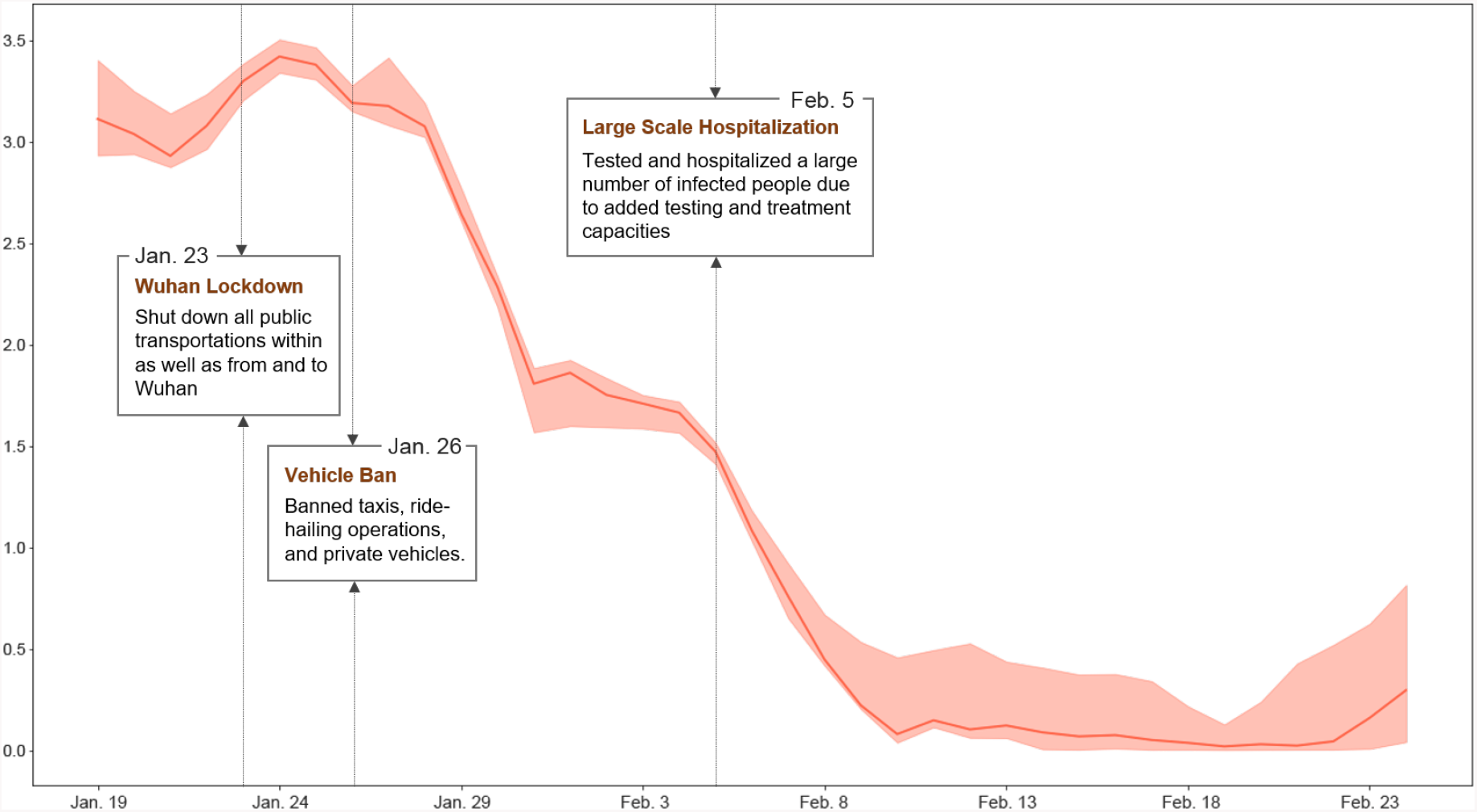
Effective Reproduction Number in Wuhan. The figure presents the evolution of the effective reproduction number in Wuhan, along with major government measures to control the outbreak. The shaded area represents the 95% credible interval. The effective reproduction number attained its maximum on January 24, 2020, which was 3.42 [95% Cl 3.34--3.50] and became less than 1 from February 7, 2020 (0.76 [95% Cl 0.65-0.92]).

### Under-reporting Factor

A key feature of our method is an attempt to recover true numbers of infections from their respective official numbers reported by the government. This is done by introducing transformation functions with under-reporting factors, and calibrating them via a Bayesian estimation approach, which is discussed in detail in the Method section. Fig. 3 shows the dynamics of the under-reporting factor *a* for the period between January 19, 2020 and February 24, 2020. Note that *a* is the ratio of the official daily increased number of infected and quarantined cases to its respective true number. Like *R, a* is also estimated using a rolling-window approach and *a* of day *t* denotes the under-reporting ratio over the time window of [*t, t* + 10].

**Fig. 3.**
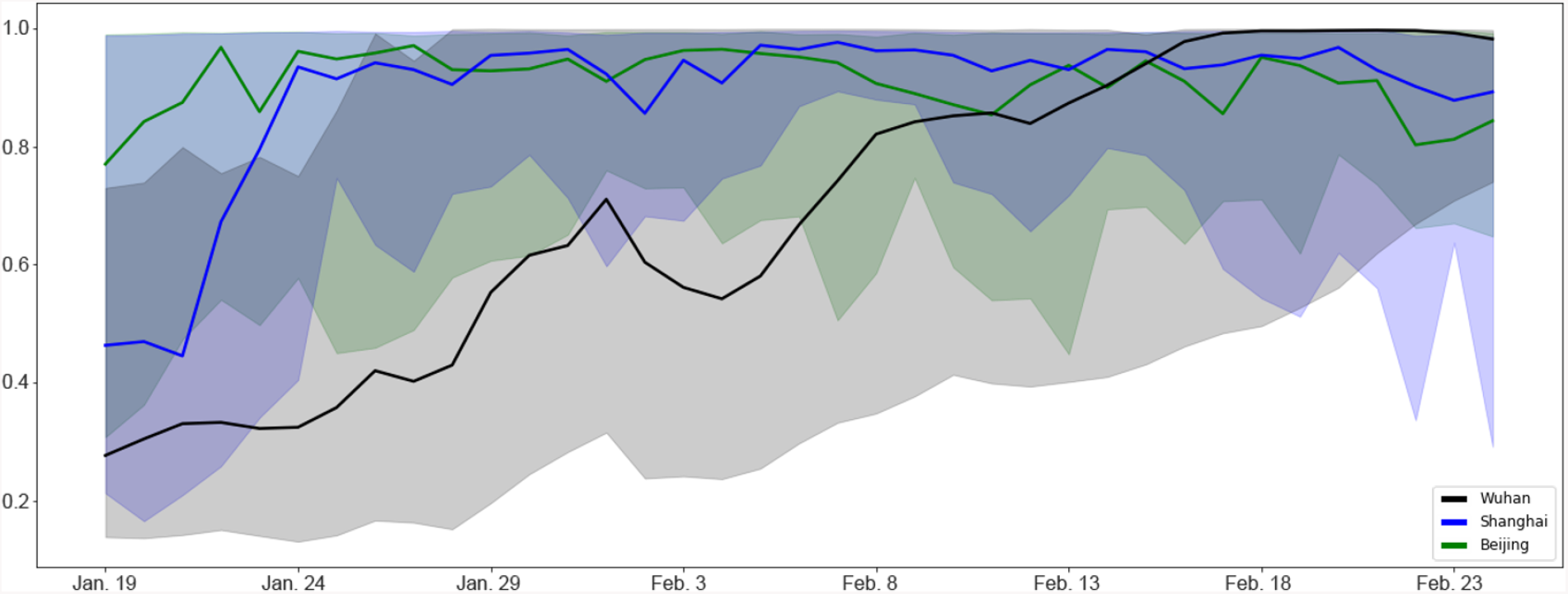
Under-reporting Factor *a* of Wuhan, Shanghai, and Beijing. The figure presents the dynamics of the under-reporting factor *a* of Wuhan (solid black line), in comparison to that of Shanghai (solid blue line) and Beijing (solid green line). The shaded areas represent the 95% credible interval.

Fig. 3 plots *a* of Wuhan in a solid black line, with the shaded area representing the 95% credible interval. As shown, *a* of January 19, 2020 was 0.28 [95% CI 0.14–0.73], indicating that official daily increased numbers of infected and quarantined cases over the window of January 19, 2020 to January 29, 2020 were on average 28% of their respective true numbers. The under-reporting factor of Wuhan gradually increased over time. For example, the under-reporting ratio over the window of January 29, 2020 to February 8, 2020 was 0.55 [95% CI N0.20–0.99] and that over the window of February 15, 2020 to February 25, 2020 was 0.94 [95% CI 0.43–0.99]. The evolution of *a* in Wuhan is in alignment with the reality. Due to insufficient testing and treatment capacities at the beginning of the observation period, many infected people were not tested or hospitalized hence not on government statistics. Through the addition of testing and treatment facilities, more infected people got tested and hospitalized, thereby increasing the under-reporting factor. Fig. 3 also presents the under-reporting factor of Shanghai and Beijing in a solid blue line and a solid green line, respectively. Clearly, all three cities underreported the actual number of quarantined cases at the beginning. While Shanghai and Beijing improved the reporting accuracy quickly, Wuhan did not catch up until the end of period. This result is consistent with the fact that Wuhan experienced explosive number of COVID-19 infections in contrast to the other two cities. But it did not have sufficient medical resources and hospital capacity to test and treat all the infected cases. The discrepancies bet.ween true and official numbers of infections in Fig. 3 imply that a data transformation approach, such as the one proposed in this paper, is necessary before estimating the epidemiological parameters of the COVID-19 outbreak in Wuhan.

### Effects of Interventions

We analyze the effects of two major government interventions on the spread of COVID-19 in Wuhan: transportation suspension and large scale hospitalization. On January 23, 2020, the municipal government suspended all public transportation services, including buses, ferries, and subways. On January 26, 2020, the government further banned taxis, ride-hailing, and private vehicle operations. These two measures constitute the intervention of transportation suspension in Wuhan, which essentially shut down the transportations in the city. It is noted that our analysis here is distinct from the study in Chinazzi et al. ^7^: the former analyzes the effect of transportation suspension in Wuhan on the spread of COVID-19 in the city, while the latter studies the effect of the transportation restrictions from and to Wuhan on the spread of COVID-19 nationally and internationally.

To evaluate the effect of transportation suspension, we focused on the period between January 26, 2020 and February 4, 2020, during which the only major intervention is transportation suspension. Fig. 4 (A) plots the true cumulative number of infected cases estimated by our method during the period in a solid blue line, with the shaded area representing the 95% credible interval. Note that these numbers reflect the spread of COVID-19 in Wuhan under the intervention of transportation suspension. To simulate the hypothetical scenario that this intervention were not imposed, we used the SIQR model parameters estimated by our method for the window period between January 21, 2020 and January 26, 2020 when no intervention effect from transportation suspension was involved. We then ran the SIQR model for the evaluation period, with the estimated infective number on January 26, 2020 as the initial state, and computed the cumulative numbers of infected cases without the intervention. Fig. 4 (A) plots the computed cumulative numbers of infected cases without the intervention (dotted green line). In particular, by February 4, 2020, in the absence of the intervention, the number of infections would be expected to climb up to 117,842 [95% CI 55,098–238,212]. Using this number as the benchmark, the number of infections saved by the intervention during the evaluation period was 33,719, resulting in 29% reduction from the scenario of no intervention. Wuhan is a metropolitan area with an average of 8 million passengers using the city’s public and private transportations daily^15,16^. Shutting down the transportations reduced the average contact rate among the city residents. As a result, the adequate contact rate *β* was decreased^17^ and the number of infections was reduced. See also the Method section for the parameter details.

**Fig. 4.**
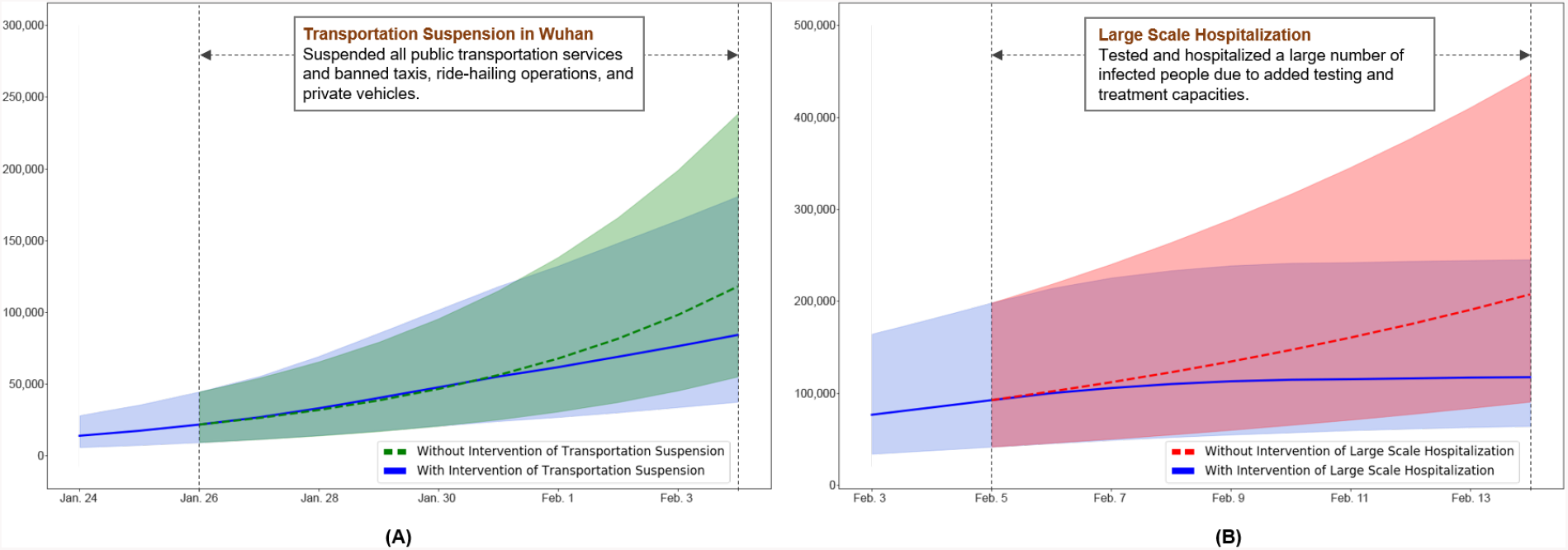
Effects of Interventions. **(A)** We evaluate the effect of transportation suspension in Wuhan for the period between January 26, 2020 and February 4, 2020. The estimated true cumulative number of infected cases under the intervention is plotted in a solid blue line whereas the computed cumulative number of infections without the intervention is in a dotted green line. The shaded areas represent the 95% credible interval. **(B)** We evaluate the effect of large scale hospitalization for the period between February 5, 2020 and February 14, 2020. The estimated true cumulative number of infected cases under the intervention is plotted in a solid blue line and the hypothetical cumulative number of infections without the intervention is in a dotted red line. The shaded areas represent the 95% credible interval.

The other intervention is large scale hospitalization started on February 5, 2020. To investigate the effect of the intervention, we studied the period between February 5, 2020 and February 14, 2020, within which large scale hospitalization is the only major intervention. To quantitatively evaluate what would have occurred without the intervention, we used the SIQR parameters estimated by our method for the window between January 31, 2020 and February 5, 2020 to exclude any intervention effect of large scale hospitalization. We then ran the SIQR model to compute the hypothetical trajectory of the cumulative numbers of infected cases for the evaluation period, with the estimated number of infections on February 5, 2020 as the initial state. In Fig. 4 (B), the trajectories are plotted in a dotted red line, in comparison to the estimated true cumulative numbers of infected cases under the intervention (solid blue line), with the shaded areas representing the 95% credible interval. During the evaluation period, if the intervention of large scale hospitalization had not been imposed, the number of infections would be expected to be 207,123 [95% CI 90,436–446,456] by February 14, 2020. With this benchmark number, the number of infections that had been prevented was 90,072, giving 43% reduction from the scenario of no intervention. The implementation of this intervention relied on the establishment and operation of two emergency specialty field hospitals, the Vulcan Mountain Hospital and the Thunder Mountain Hospital, sixteen temporary makeshift hospitals ^18^, as well as the Fire Eye Lab that enabled massive nucleic acid detection^14^. These hospitals in total had roughly 15,000 beds, which significantly increased the quarantine and treatment capacity of the public health system^19^. The added testing and treatment capacities due to the intervention allowed more timely identification and isolation of infected people, thereby reducing the number of infections.

## Discussion

Our study aims to characterize the evolution of the COVID-19 outbreak in Wuhan and reveal the effects of major government interventions on its spread. The underlying challenge in studying the pandemic dynamics lies in the discrepancy between the officially reported number of infected cases in Wuhan and the actual number of infections, in together with the lack of reliable data sources after the city’s complete lockdown (e.g., most existing work focuses on static estimation before the lockdown on January 23, 2020 and often relies on exported case numbers^4,5,10^). To address the data discrepancy issue, we employ a straight-forward yet effective data transformation approach under a Bayesian dynamic epidemic modeling framework, which leads to important implications in understanding the evolution of Wuhan’s outbreak. First, using prior literature knowledge on COVID-19, we adjust for the reported data to estimate and gauge the actual outbreak sizes, which is shown to be substantially larger than those from official reports particularly in early periods. Second, taking into account the adjusted numbers, the resulting trajectory for effective reproduction numbers serves as more accurate reflection of disease spread trends and the temporal changes in response to official intervention policies. Third, our study results are crucially equipped with under-reporting factors that, to some extent, reflect the difficulty level in recording the actual infective numbers and the stress of COVID-19 on medical resources. In particular, by comparison with two other major cities in China, our results from the under-reporting factors are in alignment with the reality that Wuhan as the epicenter experienced the longest periods of high stress on health care system while the numbers outside Wuhan tend to be generally trustworthy at smaller outbreak scale with better medical preparedness.

Although our study uncovers some convincing approximation on the dynamic progression patterns of COVID-19 in Wuhan, there remain some limitations. Here we assume that all recovered patients become totally immune to the novel coronavirus infection. If recovered patients are still susceptible, an extension from SIQR to SIQRS (that is, Susceptible-Infective-Quarantined-Removed-Susceptible) may be employed, while the general framework of our method remains largely applicable. In addition, the removed compartment in our model contains both death and cured cases, which prevents us from estimating the time-varying case fatality rates. Consequently, our assessment of large scale hospitalization does not reflect its effectiveness in death toll reduction, although literature has shown that promptly hospitalizing infected people could reduce the fatality rate for older adults and even for those with mild symptoms^20,21,22,23,24^. Future studies may investigate the trajectory of fatality rates by treating death and cured cases separately.

In summary, our finding provides a quantitative illustration that the scale of infection size in Wuhan can be multi-fold higher than officially reported numbers and partially explains the excessive stress experienced by frontline medical workers despite seemingly modest case number increases reported during late January of 2020. This work thus gives a cautionary tale for drawing immediate public health conclusions solely based on unadjusted official case numbers that do not necessarily give a complete overall picture for pandemic situation in outbreak early periods. In addition, by examining the temporal trajectory of effective reproduction numbers, we can clearly see the gradual control effects of COVID-19 in Wuhan soon after the implementation of city-wide lockdown and suspension of all non-essential vehicle operation to reduce the contact rate among Wuhan residents; the aggressive increase of testing and hospital capacity further brought down the effective reproduction number rapidly by shortening infectious period of positive carriers and reducing new cross-infection cases from close family and community contacts. Our study affirms the believed importance and effectiveness of imposing tight non-essential travel restrictions (which may also include, e.g., the shelter-in-place and stay-at-home orders) early on, as well as swiftly addressing the testing shortage issues and avoiding hospital overcrowding for effective mitigation of COVID-19 community spread.

## Method

### Data

We obtained data about the COVID-19 outbreak in Wuhan from official reports released by the Chinese Center for Disease Control and Prevention (CCDC) between January 18, 2020 and March 5, 2020. CCDC provides daily cumulative number of infected cases and removed cases (i.e., recovery and death). Let 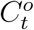 denote the cumulative number of infected cases by day *t* and 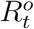 be the cumulative number of removed cases by day *t*, both officially released by CCDC. Assuming that all the officially confirmed infections have been effectively quarantined (e.g., hospitalized), we have

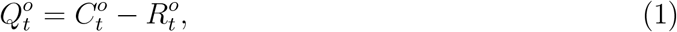

where 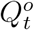 is the official number of actively infected and quarantined cases by day *t*.

It is worth noting that daily number of newly infected cases dramatically increased to 13,436 on February 12, 2020 from 1,104 the day before, according to CCDC. This surge was attributed to the change of government criteria for confirming infections. Before February 12, 2020, only those tested positive by test kits were considered as infected. Starting from February 12, 2020, an infection was confirmed either based on positive testing result or through clinical diagnosis using computed tomography (CT) scans. As a result, suspected infections by CT scans before February 12, 2020 were relabeled as confirmed infections on February 12, 2020. It is therefore necessary to adjust the number of newly infected cases on February 12, 2020 (i.e., 13,436) by reallocating this number to days prior to and including February 12, 2020, proportional to the number of daily suspected cases in these days.

### Method Overview

We assume that the diffusion of COVID-19 in Wuhan follows an epidemic model whose underlying time-dependent state variables 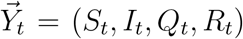 are from a dynamic system with system parameters Θ_*H*_ = (*β, µ, γ*). These state variables and system parameters are summarized in Table 1; their meanings and the epidemic model will be elaborated in the next subsection. In particular, *Q*_*t*_ represents the number of actively infected and quarantined cases by day *t* and *R*_*t*_ represents the cumulative number of removed cases by day *t*.

**Table 1:** Notation for the SIQR model.

| Notation | Description |
| --- | --- |
| $N$ | population size |
| $S_t$ | number of susceptible cases at day $t$ |
| $I_t$ | number of actively infected but not quarantined cases by day $t$ |
| $Q_t$ | number of actively infected and quarantined cases by day $t$ |
| $R_t$ | cumulative number of removed cases by day $t$ |
| $\beta$ | adequate contact rate |
| $\mu$ | rate at which an infected case gets quarantined |
| $\gamma$ | rate at which a quarantined case becomes removed |

Ideally, we can obtain data about actual diffusion of COVID-19 over time. That is, ideally, we can have stochastically realized true values of *Q*_*t*_ and *R*_*t*_ for *t* = 1, 2, 3, …, denoted as 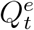and 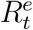. In general, if the realized true values of all state variables were known, we could estimate system parameters Θ_*H*_ using well-developed statistical methods (e.g., ^25,26,27^ from frequentist perspectives). In reality, we only observe a subset of state variables with their officially reported numbers 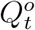 and 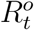. Due to the under-reporting problem, these official numbers, 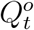 and 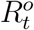, could be much lower than 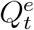 and 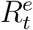, respectively. As a result, directly applying an existing method 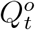, and 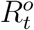 may not generate or reliably uncover the epidemiological parameters of COVID-19. To address this issue, we propose transformation functions that aim to recover 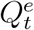 and 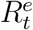 from observed 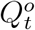 and 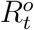 with some (unknown) transformation parameters Θ_*f*_.

With the aforementioned framework, we need to estimate parameters Θ_*H*_ and Θ_*f*_. Instead of using the frequentest approaches (such as maximum likelihood estimation or MLE), we develop an Bayesian approach for our problem because of the following considerations. First, the Bayesian approach allows us to incorporate existing knowledge on COVID-19 to give a guided estimation of Θ_*H*_ through well-informed prior selection, while the MLE approach would have to largely ignore the valuable information from prior literature. Second, the posterior distribution, given our proposed modeling strategy and prior, has clear interpretation and can provide straightforward uncertainty quantification. To our knowledge, the MLE approach for our specified model settings has no well-developed inference theory for the estimators. Third, from a practical perspective, our Bayesian sampling scheme (described in the subsection of Parameter Estimation) for the posterior distributions is straightforward to derive and implement, while the MLE estimator is more computationally involved and difficult to obtain.

For explicit overview summary, we include all the essential components of our Bayesian modeling scheme for an epidemic model with transformation functions proposed above in Fig. 5, whose technical details will be described in the following subsections.

**Fig. 5.**
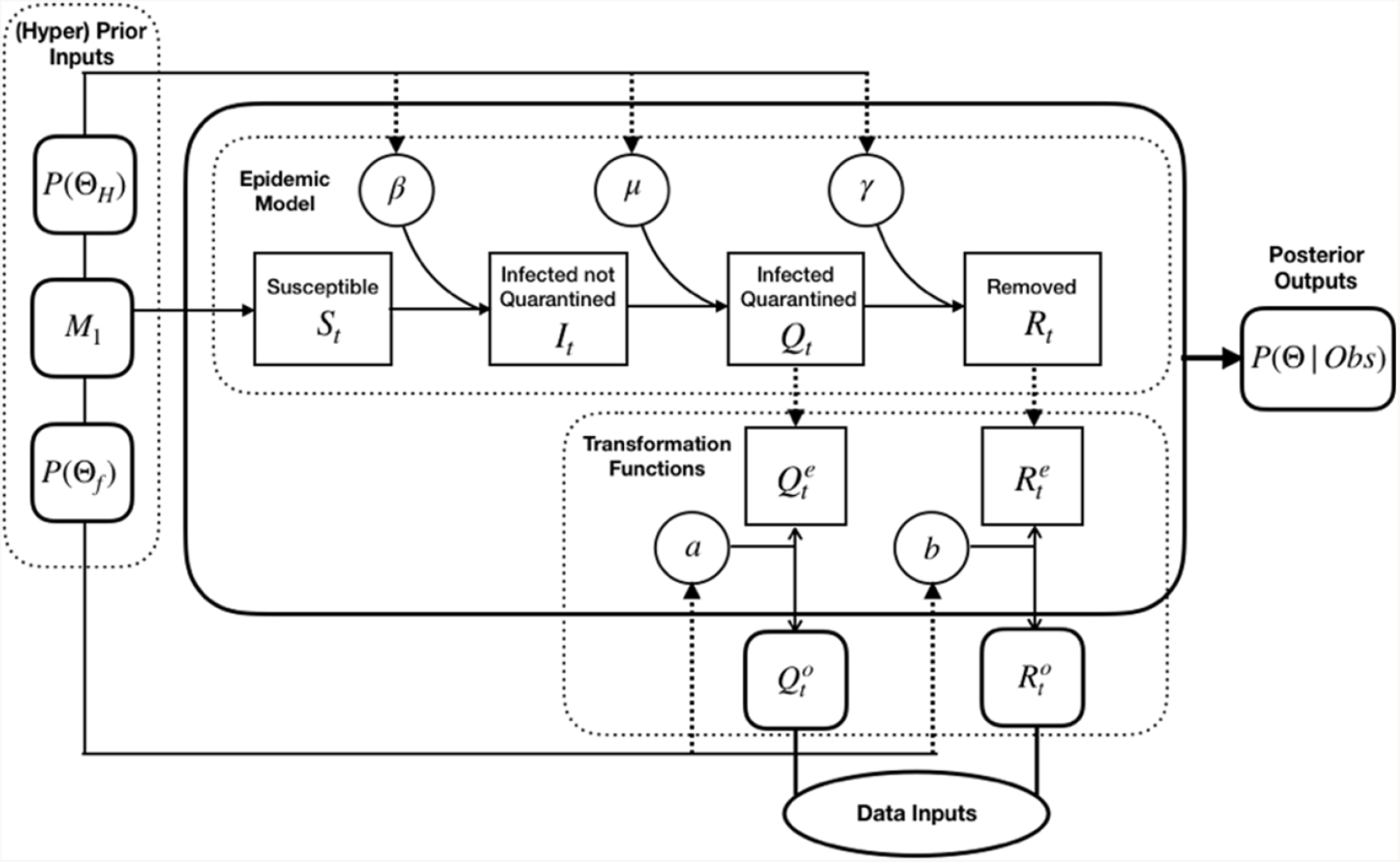
Bayesian Estimation Scheme for an Epidemic Model with Transformation Functions.

### Epidemic Model

Recent evidences have shown that non-symptomatic infected cases and infected cases in their latent period can spread COVID-19 with high efficiency, e.g., Chang et al. ^13^. In alignment with these findings, we adopt a Susceptible-Infective-Quarantined-Removed (SIQR) compartmental model to characterize the diffusion of COVID-19^28^. The susceptible compartment of the model consists of those who can be infected. The infective compartment is composed of those who are actively infected but not quarantined, with or without symptoms. Those who are actively infected and quarantined are in the quarantined compartment. The removed compartment consists of those who recover or die from the disease. The state variables of the epidemic model, *S*_*t*_, *I*_*t*_, *Q*_*t*_, and *R*_*t*_, are defined in Table 1, and the population size *N* = *S*_*t*_ + *I*_*t*_ + *Q*_*t*_ + *R*_*t*_ ^29^. The SIQR model is defined using the following ordinary differential equations (ODE):

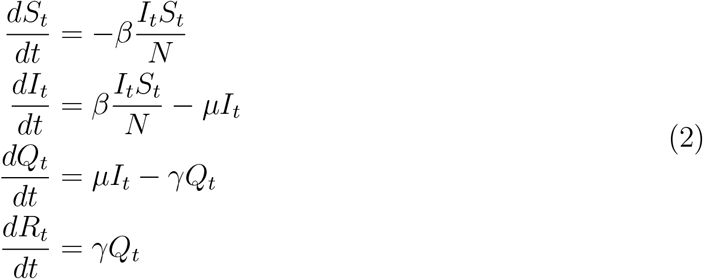

In these ODEs, *β* is the adequate contact rate, where adequate contacts refer to contacts sufficient for transmission^30^. *µ* is the rate at which an infected case gets quarantined, and *γ* is the rate at which a quarantined case becomes removed. In the SIQR model, the effective reproduction number *R* and the cumulative number *M*_*t*_ of infected cases by day *t* are given by ^28,31^

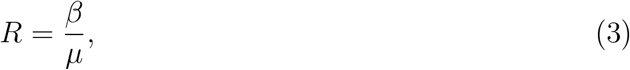

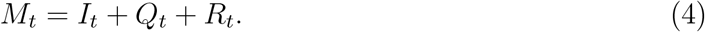

### Transformation Functions

Let 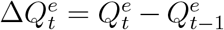 be the true daily increased number of infected and quarantined cases at day *t*. Similarly, let 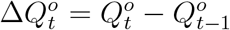 be the officially reported daily increased number of infected and quarantined cases at day *t*, i.e., the official counterpart of 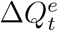. Due to the underreporting problem, 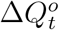 tends to be smaller than 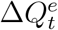. Assuming that the daily increased number of infected and quarantined cases is underreported in a consistent manner within a short time window, we model the relationship between 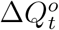 and 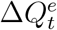 as

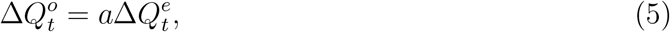

where 0 < *a* ≤ 1 is the underreporting factor of quarantined cases. Clearly, the greater the value of *a*, the closer the official number 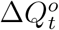 to the true number 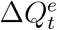. By (5), we derive 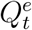 as

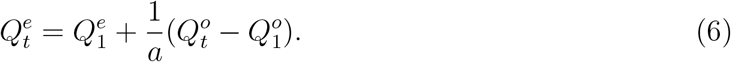

Let 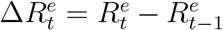 denote the true daily increased number of removed cases at day *t* and 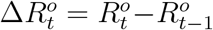 be the official counterpart of 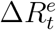. Similarly, we model the relationship between 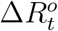 and 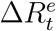 in a short time window as

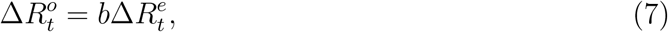

where 0 < *b* ≤ 1 is the underreporting factor of removed cases. By (7), we derive 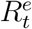 as

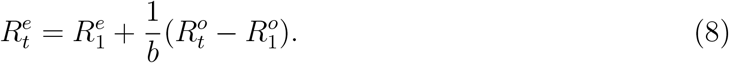

Although both (5) and (7) have seemingly simple formats, they catch the relationships between true and official numbers well as demonstrated in our empirical analysis. Moreover, our method is flexible and using other alternative functional forms to model the relationships between true and official numbers dose not affect the general framework of our method.

### Parameter Estimation

Having defined the general framework of the epidemic model with transformation functions, we next show how to learn its associated parameters, Θ = Θ_*H*_ *∪* Θ_*f*_ = (*β, µ, γ, a, b*). Specifically, we impose a prior distribution *P* (Θ) on Θ by resorting to existing knowledge on COVID-19 and obtain the posterior distribution of Θ given the reported discrete trajectory of official numbers 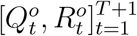, where the short time window is from *t* = 1 to *t* = *T* + 1. Accordingly, we obtain the unnormalized posterior distribution 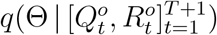 as

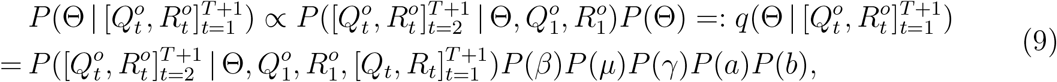

where expanding the condition set in the last equality from 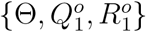 to 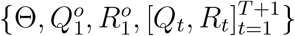 adds no new information because given 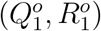 and Θ, 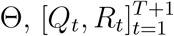 can be deterministically derived using the SIQR model (with initial state variables explained in the next subsection). Also, we use independent priors *P* (Θ) = *P* (*β*)*P* (*µ*)*P* (*γ*)*P* (*a*)*P* (*b*).

To find appropriate priors, we note from Sun et al. ^32^ that the median incubation period of COVID-19 is estimated to be 4.5 days with interquartile range (IQR) 3.0-5.5 days, and the median delay between symptom onset and seeking care is 2 days with IQR 0-5 days in mainland China after January 18, 2020, the starting date of our analysis. Therefore the infectious period of COVID-19 ranges from 3 to 10.5 days. Accordingly, we set parameter *µ* to be uniformly distributed over 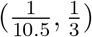.

In addition, to the best of our knowledge, there is no literature on the duration from quarantine to removal for COVID-19 infected cases. Therefore, we collect data about 32 death cases and 22 cured cases in Wuhan from local newspapers. Details of these cases are given in Supplementary Table 2. Among the death cases, the minimum duration of hospitalization is 1 day and the maximum is 40 days. The range of hospitalization for cured cases is from 6 to 30 days. For COVID-19 infected cases in Wuhan, the percentage ratios of death and cure are 5.8% and 94.2%, respectively^33^. Accordingly, we roughly estimate the duration from quarantine to removal in the SIQR model to have range from 5.7 to 30.6 days using weighted averages, and we set parameter *γ* to be uniformly distributed over 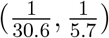. Non-informative flat priors are adopted for the rest parameters so that (10) in the following summarizes our empirical prior settings.

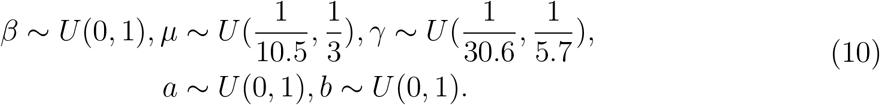

We further assume that for *t* = 2, *&, T* + 1, true numbers 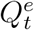 and 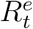 follow a Poisson distributions with means *Q*_*t*_ and *R*_*t*_, respectively. In together with the relation between true numbers 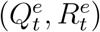 and official numbers 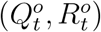 from (6) and (8), we use

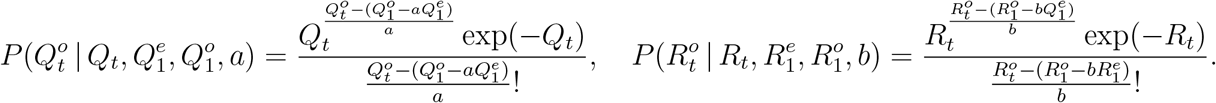

By the following conditional independence, we compute the unnormalized posterior through

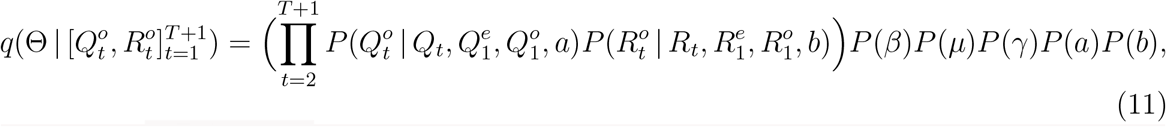

where 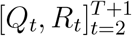 are generated from model (2) given Θ, 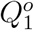 and 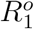. Following the Metropolis–Hastings algorithm (e.g., Geyer and Thompson ^34^), we obtain the estimation of parameters by employing the Markov chain Monte Carlo (MCMC) sampling from (11). Specifically, suppose Θ_(*k-*1)_ is the current state of the Markov chain, and let *J* (Θ | Θ_(*k-*1)_ be the jumping distribution chosen to be independent normals with mean Θ_(*k-*1)_ an elementwise variance *c*^2^, where *c* is a scale parameter for rejection rate adjustment. The MCMC sampling proposes Θ^*∗*^ from *J* (Θ | Θ_(*k-*1)_) and computes

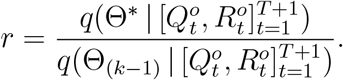

The next state is then set to be Θ_(*k*)_ = Θ^*∗*^*Z* + Θ_(*k-*1)_(1 *-Z*), where *Z* has Bernoulli distribution with probability parameter min(1, *r*). If {Θ^(*l*)^}_*l*=1,…,*K*_ is the MCMC sample obtained after a “burn-in” period, the posterior mean estimator is approximated as 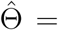 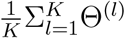.

### Dynamic Parameter Estimation Over Time

Since the Chinese government responds with evolving containment and mitigation actions towards the development of COVID-19, to obtain updated information on the parameters Θ, we adopt a rolling window approach to estimate Θ for each short time period [*t, t*+1, …, *t*+*T*], where the window size is *T* days and *t* = 1, 2, 3,. In this study, we use a 10-day time window, i.e., *T* = 10; also the first day with *t* = 1 in our analysis corresponds to January 18, 2020. For each time period starting at *t*, we denote Θ_*t*_ = Θ as the parameters of interests. The posterior 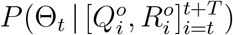 is learned using the reported discrete trajectory of official numbers 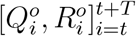 in the window of [*t, t* + 1, …, *t* + *T*]. While the trajectory of official numbers 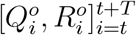 can be observed, we need to set the initial true numbers 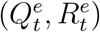.

Besides, noting that to complete our Bayesian estimation scheme, we need to set initial values for the epidemic model. Correspondingly, for *t* = 1, we set (*Q*_1_, *R*_1_) as 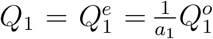, and 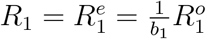 which implies that

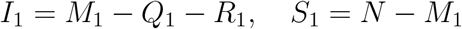

where *a*_1_ and *b*_1_ are the corresponding under-reporting factors for the time period of [1, 2, …, *T* + 1], and *M*_1_ represents the true cumulative number of infections by day 1 or January 18, 2020. Using the number of infected cases exported from Wuhan internationally, Imai et al. ^12^ estimate that the cumulative number of infections in Wuhan by January 18, 2020 is 4,000 with a 95% confidence interval [1,700-7,800] in the baseline scenario. Additionally, to account for 2 million people leaving Wuhan due to Wuhan lockdown on January 23, 2020, we set the population size *N* to be 11 million (i.e., regular population size in Wuhan^35^) before January 23, 2020 and adjust it to 9 million after January 23, 2020^36^. With the above setting and the observed official numbers 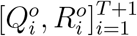, we can estimate parameters Θ_1_ = (*β*_1_, *µ*_1_, *γ*_1_, *a*_1_, *b*_1_) and compute 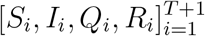. Subsequently, the computed (*S*_2_, *I*_2_, *Q*_2_, *R*_2_) can serve as the initial values for the second time window [2, 3, …, *T* +2], and we continue this strategy as the rolling window moves forward. Consequently, the proposed dynamic parameter estimation procedure is expected to track the trend of the epidemiological parameters of COVID-19 and dynamically assesses temporally evolving situations.

## Data Availability

Data are available upon reasonable request.

## Acknowledgements

We thank Huiran Li for her help with literature review and data processing.

## Supplementary Tables

**Supplementary Table 1.** Estimated true cumulative number of infected cases in Wuhan. We estimated the true cumulative number of infections in Wuhan or the outbreak size by each day for the period between January 19, 2020 and March 5, 2020. The input to our method is the cumulative number of infections in Wuhan by January 18, 2020 estimated in Imai et al. ^1^, whose baseline estimate is 4,000 with a 95% confidence interval [1,700–7,800].

| Date | Estimated True Cumulative Number<br>of Infected Cases | Lower Bound<br>95% CI | Upper Bound<br>95% CI |
| --- | --- | --- | --- |
| 1/19/20 | 4,932 | 2,045 | 9,579 |
| 1/20/20 | 6,120 | 2,552 | 11,931 |
| 1/21/20 | 7,510 | 3,166 | 14,798 |
| 1/22/20 | 9,151 | 3,875 | 18,095 |
| 1/23/20 | 11,239 | 4,794 | 22,372 |
| 1/24/20 | 13,921 | 5,956 | 28,064 |
| 1/25/20 | 17,394 | 7,501 | 35,474 |
| 1/26/20 | 21,665 | 9,375 | 44,570 |
| 1/27/20 | 26,785 | 11,611 | 55,254 |
| 1/28/20 | 33,034 | 14,293 | 69,331 |
| 1/29/20 | 40,244 | 17,466 | 85,412 |
| 1/30/20 | 47,671 | 20,885 | 101,725 |
| 1/31/20 | 55,146 | 24,093 | 117,964 |
| 2/1/20 | 61,674 | 26,998 | 132,278 |
| 2/2/20 | 68,939 | 30,248 | 148,255 |
| 2/3/20 | 76,279 | 33,780 | 164,095 |
| 2/4/20 | 84,122 | 37,568 | 180,687 |
| 2/5/20 | 92,158 | 41,492 | 197,990 |
| 2/6/20 | 99,676 | 45,329 | 213,730 |
| 2/7/20 | 105,347 | 48,842 | 225,268 |
| 2/8/20 | 109,658 | 51,955 | 233,031 |
| 2/9/20 | 112,622 | 54,666 | 238,526 |
| 2/10/20 | 114,270 | 57,075 | 241,236 |
| 2/11/20 | 114,900 | 59,480 | 241,997 |
| 2/12/20 | 115,723 | 61,262 | 243,384 |
| 2/13/20 | 116,636 | 62,810 | 244,251 |
| 2/14/20 | 117,050 | 64,004 | 245,014 |
| 2/15/20 | 117,862 | 65,038 | 245,292 |
| 2/16/20 | 118,302 | 65,956 | 245,454 |
| 2/17/20 | 118,920 | 66,697 | 245,748 |
| 2/18/20 | 119,135 | 67,211 | 245,877 |
| 2/19/20 | 119,299 | 67,422 | 246,021 |
| 2/20/20 | 119,571 | 67,433 | 246,232 |
| 2/21/20 | 120,098 | 67,578 | 246,622 |
| 2/22/20 | 120,787 | 67,834 | 247,289 |
| 2/23/20 | 120,953 | 68,080 | 248,483 |
| 2/24/20 | 121,651 | 68,295 | 249,819 |
| 2/25/20 | 122,481 | 68,508 | 251,638 |
| 2/26/20 | 122,784 | 68,692 | 253,385 |
| 2/27/20 | 123,063 | 68,853 | 255,063 |
| 2/28/20 | 123,320 | 68,994 | 256,674 |
| 2/29/20 | 123,557 | 69,119 | 258,221 |
| 3/1/20 | 123,777 | 69,223 | 259,708 |
| 3/2/20 | 123,980 | 69,314 | 261,141 |
| 3/3/20 | 124,168 | 69,393 | 262,523 |
| 3/4/20 | 124,344 | 69,465 | 263,851 |
| 3/5/20 | 124,506 | 69,526 | 265,113 |

**Supplementary Table 2.** Death and cured COVID-19 cases collected from Wuhan local news. This table presents the details of each death or cured COVID-19 case we collected from Wuhan local news^2,3,4,5,6,7,8^. For cases 37-54, the local news^8^ only provide overall descriptive statistics.

| Case Number | Discharge Status | Sex | Age | Hospitalization Date | Discharge Date | Days Between Hospitalization and Discharge |  |
| --- | --- | --- | --- | --- | --- | --- | --- |
| 1 | Death | M | 61 | 1/27/20 | 2/9/20 | 13 |  |
| 2 | Death | M | 69 | 1/3/20 | 1/15/20 | 12 |  |
| 3 | Death | M | 89 | 1/8/20 | 1/18/20 | 10 |  |
| 4 | Death | M | 89 | 1/13/20 | 1/19/20 | 6 |  |
| 5 | Death | M | 66 | 1/16/20 | 1/20/20 | 4 |  |
| 6 | Death | M | 75 | 1/11/20 | 1/20/20 | 9 |  |
| 7 | Death | F | 48 | 12/11/19 | 1/20/20 | 40 |  |
| 8 | Death | M | 82 | 1/14/20 | 1/21/20 | 7 |  |
| 9 | Death | M | 66 | 12/31/19 | 1/21/20 | 21 |  |
| 10 | Death | M | 81 | 1/18/20 | 1/22/20 | 4 |  |
| 11 | Death | F | 82 | 1/6/20 | 1/22/20 | 16 |  |
| 12 | Death | M | 65 | 1/11/20 | 1/21/20 | 10 |  |
| 13 | Death | F | 80 | 1/18/20 | 1/22/20 | 4 |  |
| 14 | Death | M | 53 | 1/13/20 | 1/21/20 | 8 |  |
| 15 | Death | M | 86 | 1/9/20 | 1/21/20 | 12 |  |
| 16 | Death | F | 70 | 1/13/20 | 1/21/20 | 8 |  |
| 17 | Death | M | 84 | 1/9/20 | 1/22/20 | 13 |  |
| 18 | Death | M | 70 | 1/19/20 | 1/23/20 | 4 |  |
| 19 | Death | F | 76 | 1/18/20 | 1/24/20 | 6 |  |
| 20 | Death | M | 72 | 1/18/20 | 1/23/20 | 5 |  |
| 21 | Death | M | 79 | 1/17/20 | 1/24/20 | 7 |  |
| 22 | Death | M | 55 | 1/19/20 | 1/24/20 | 5 |  |
| 23 | Death | M | 87 | 1/19/20 | 1/23/20 | 4 |  |
| 24 | Death | F | 66 | 1/19/20 | 1/21/20 | 2 |  |
| 25 | Death | M | 58 | 1/18/20 | 1/24/20 | 6 |  |
| 26 | Death | M | 66 | 1/11/20 | 1/21/20 | 10 |  |
| 27 | Death | M | 78 | 1/23/20 | 1/24/20 | 1 |  |
| 28 | Death | M | 65 | 1/16/20 | 1/23/20 | 7 |  |
| 29 | Death | M | 67 | 1/15/20 | 1/24/20 | 9 |  |
| 30 | Death | M | 58 | 1/1/20 | 1/23/20 | 22 |  |
| 31 | Death | F | 67 | 1/12/20 | 1/23/20 | 11 |  |
| 32 | Death | F | 82 | 1/17/20 | 1/23/20 | 6 |  |
| 33 | Cured | F | 52 | 1/21/20 | 2/4/20 | 14 |  |
| 34 | Cured | M | 97 | 2/5/20 | 2/22/20 | 17 |  |
| 35 | Cured | F | 97 | 1/29/20 | 2/28/20 | 30 |  |
| 36 | Cured | F | - | 1/19/20 | 1/30/20 | 11 |  |
| 37-54 | Cured | - | - | - | - | Mean | 12 |
|  |  |  |  |  |  | Min | 6 |
|  |  |  |  |  |  | Max | 18 |

